# Loss to follow-up correction increased mortality estimates in HIV-positive people on antiretroviral therapy in Mozambique

**DOI:** 10.1101/2020.08.04.20167155

**Authors:** Nanina Anderegg, Jonas Hector, Laura F Jefferys, Juan Burgos-Soto, Michael A Hobbins, Jochen Ehmer, Lukas Meier, Marloes H Maathuis, Matthias Egger

**Author notes:** <u>Correspondence to:</u> Nanina Anderegg, MSc, Institute of Social and Preventive Medicine, University of Bern, Bern, Switzerland.

## Abstract

**Objectives:** People living with HIV (PLWH) on antiretroviral therapy (ART) may be lost to follow-up (LTFU), which hampers the assessment of outcomes. We estimated mortality for patients starting ART in a rural region in sub-Saharan Africa and examined risk factors for death, correcting for LTFU.

**Study design and setting:** We analysed data from Ancuabe, Mozambique, where patients LTFU are traced by phone and home visits. We used cumulative incidence functions to estimate mortality and LTFU. To correct for LTFU, we revised outcomes based on tracing data using different inverse probability weights (maximum likelihood [ML], Ridge regression or Bayesian model averaging [BMA]). We fitted competing risk models to identify risk factors for death and LTFU.

**Results:** Analyses included 4492 patients; during 8152 person-years of follow-up, 486 patients died, 2375 were LTFU, 752 were traced, and 603 were found. At 4 years after starting ART, observed mortality was 11.9% (95% CI 10.9-13.0) but 23.5% (19.8- 28.0), 21.6% (18.7-25.0) and 23.3% (19.7- 27.6) after correction with ML, Ridge and BMA weights, respectively. Risk factors for death included male sex, lower CD4 cell counts and more advanced clinical stage.

**Conclusion:** In ART programmes with substantial LTFU, mortality estimates need to take LTFU into account.

**What is new?**

Key findings

- In an antiretroviral therapy (ART) programme in Mozambique, correction of survival estimates using inverse probability weighting (IPW) of outcomes in patients lost to follow-up led to substantially higher cumulative mortality.
- Correction also had an impact on some estimates of associations with risk factors for death.

What this adds to what is known?

- This study provides new insights into the use of different IPW weights for correction of LTFU in ART programmes in sub-Saharan Africa, including maximum likelihood weights, weights from Ridge regression or Bayesian model averaging.
- Results were generally similar, independently of the type of weight used. If all important covariates describing differences between patients traced and not traced are collected, an IPW approach using any of the three parameter estimation methods can be used to correct mortality for true outcomes in patients LTFU.

What is the implication and what should change now?

- ART programmes with high rates of LTFU should trace patients LTFU to bring them back into care and correct programme-level estimates of mortality.

## 1. Introduction

Mortality in people living with HIV (PLWH) on antiretroviral therapy (ART) is an essential indicator of ART programmes’ effectiveness [1,2]. In sub-Saharan Africa and elsewhere, many PLWH who started ART are lost to follow-up (LTFU), which makes mortality estimation challenging [3]. Patients LTFU usually experience higher mortality than those remaining in care, but their deaths are not recorded [4–8]. If these deaths are ignored, for example, by merely censoring follow-up time of patients LTFU at their last clinic visit, overall programme-level mortality will be underestimated.

Several correction methods to reduce this bias have been proposed [9–15]. These methods often rely on vital status information ascertained for a sample of patients LTFU, which is obtained by tracing patients LTFU by phone or home visits, or by data linkage with vital registries [4,6,7,16]. One approach treats the sample of ascertained subjects as representative of all patients LTFU. Patients LTFU with no vital status information are excluded from analyses, while those whose vital status could be determined receive weights proportional to the inverse of their predicted probabilities of ascertainment [14,15,17].

Mozambique is among the countries most affected by HIV worldwide [18,19], and poor retention is a major challenge for ART programmes [20,21]. We aimed to estimate mortality and LTFU in PLWH starting ART in rural Mozambique and to identify risk factors for death and LTFU. Using tracing data, we used three different inverse probability weights to correct for true outcomes in patients LTFU and examined the sensitivity of estimates to different weights.

## 2. Methods

### 2.1 Setting and data

We analysed data from two ART clinics from the rural Ancuabe district in Mozambique, which are part of the International epidemiology Databases to Evaluate AIDS (IeDEA) [22]. We included all adults (≥15 years) who started ART from 2005-2016. Visits for patients were scheduled monthly during this period. Patients who were part of a community ART group (CAG) were allowed to rotate their monthly visits within their group. A Visit of a member of the CAG was counted as a visit for all members of the group. Since 2014, clinic personnel traced a sample of patients who missed a follow-up visit by more than two months to bring them back to care and establish their vital status. The patients to be traced were selected by clinic staff (from monthly updated lists containing all patients ever having missed a visit and not yet returned to the clinic) and did not represent a random sample. The Ethics Committee of the Canton of Bern (150/15) and Mozambique’s National Bioethics Committee for Health (327/CNBS/15) approved the use of routine clinical data for research within the IeDEA collaboration. In IeDEA, informed consent is not required for the routine data analysed in the present study.

### 2.2 Outcomes

The outcomes of interest were death and LTFU. In uncorrected analyses, we used information on vital status from the clinical database only and defined LTFU as no clinic visit within six months before database closure. We measured follow-up time from ART start to date of death or last clinic visit. Outcomes were then corrected by incorporating information from tracing data. The outcome status of located patients was changed to “dead” or “alive” and follow-up times were revised, replacing the date of the last clinic visit with the date the patient was traced and found alive, or with the date of death. Patients who were tracked but not found continued to be classified as LTFU (see supplemental <u>Figure S1</u>).

### 2.3 Variables

Explanatory variables included the period of starting ART, sex, age, WHO clinical stage [23], and CD4 cell count, all at ART start. The period of starting ART was grouped into 2005-2013 and 2014-2016 to reflect the change in WHO guidelines on when to start ART [24,25] in 2013. Age and CD4 count were treated as continuous variables and centred. WHO stage was grouped into “I & II” and “III & IV” to separate patients with no symptoms or mild disease from those with advanced disease [23].

### 2.4 Imputation of CD4 cell counts

We used multiple imputation under the assumption of missingness at random to impute missing CD4 counts. We used an approach that is superior to standard software implementations if the primary analysis consists of non-linear models or includes interactions, both of which applies to our study [26]. We generated 50 imputed data sets and combined results by using Rubin’s rule and a method based on moments [27,28]. See supplementary <u>Text S1</u> for technical details.

### 2.5 Correction for LTFU

We did all analyses with and without correction for true outcomes in patients LTFU. In corrected analyses, we used the revised outcome status and follow-up times and assigned inverse-probability weights (*wi*) to individuals:

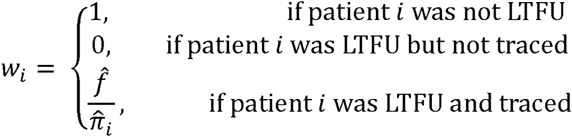

Here 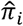 is the probability of individual *i* who was LTFU to be traced, and 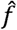 is a data-dependent factor such that weights of traced patients add up to the total number of patients LTFU. To estimate 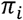 we restricted data to the subset of patients originally LTFU and modelled the odds of being traced by logistic regression. We included all explanatory variables described above. In addition, we included the duration of being LTFU (grouped into <1 year and ≥1 year) and all two-way interactions. We estimated weights with three approaches within this logistic regression framework: (i) maximum likelihood estimation (ML); (ii) penalised maximum likelihood estimation by Ridge regression (RIDGE); and (iii) Bayesian model averaging (BMA). We truncated weights at their 98^th^ percentile [29,30].

### 2.6 Competing risk regression

We calculated non-parametric cumulative incidence functions of death and LTFU. We fitted cause-specific hazard models to assess associations between patient characteristics and hazard of death and LTFU and subdistribution hazard models to evaluate associations between patient characteristics and cumulative incidence of death and LTFU [31–34]. To model the cause-specific hazards, we included all explanatory variables and two-way interactions to estimate hazard ratios (HRs) and 95% confidence intervals (CIs). Subdistribution HRs (sHRs) cannot be interpreted as rate or risk ratios [35]. However, the direction of estimated sHRs reflects the associations between covariates and the cumulative incidence of outcomes [36]. Also, statistical inference from hypothesis testing is valid, and if covariates are standardised, the sHRs reflect the relative importance of variables. We used the same model structure for the subdistribution hazard model as for the cause-specific model but standardised all variables (including categorical ones). To evaluate model fit, we examined Cox-Snell residuals, and to check the proportional hazards assumption we used Grambsch and Therneau’s goodness of fit tests [37].

### 2.7 Sensitivity analysis

In a sensitivity analysis, we fitted models to complete cases (with CD4 counts available) to assess the impact of multiple imputation of CD4 counts.

## 3. Results

<u>Table 1</u> reports patient characteristics and original and corrected outcomes for all patients and observed outcomes for traced patients. In total, 4492 patients were included; 3093 (69%) of them were female. Most of them started ART in clinical stage I & II (2558, 57%). Among all patients, 2375 (53%) were LTFU, of whom 752 (32%) were traced. Of the traced patients, 557 (74%) were found alive, 46 (6%) had died, and 149 (20%) were not found. Median time from last clinic visit to tracing was 133 days, ranging from being traced directly after a missed clinic visit to being traced 4.5 years after being lost. Most patients were traced within one year of being LTFU (617 out of 752, 82%) and only 135 (18%) were lost for longer before being traced (supplementary <u>Table S1</u>). The characteristics of patients traced were similar to all patients, and characteristics were similar after correction using tracing data (<u>Table 1</u>, supplementary <u>Table S2</u>). For 1168 (26%) of patients, the CD4 cell count was missing. Comparing patients with and without missing CD4 counts at ART initiation, those with a missing value were more likely to be female, start ART in later periods and with less advanced clinical stages and more likely to have been LTFU (supplementary <u>Table S3</u>).

**Table 1:** Baseline characteristics and outcomes at database closure or tracing of patients starting antiretroviral therapy in Ancuabe, rural Mozambique.

|  | All patients<br>(Original outcomes)<br>n=4492 |  |  | Patients lost to follow-up and traced<br>(Tracing outcomes)<br>n=752 |  |  | All patients<br>(Corrected outcomes*)<br>n=4492 |  |  |
| --- | --- | --- | --- | --- | --- | --- | --- | --- | --- |
|  | Alive | Died | Lost to follow-up | Alive | Died | Not found | Alive | Died | Lost to follow-up |
| **Total** | 1631 | 486 | 2375 | 557 | 46 | 149 | 3276 | 761 | 455 |
| Female | 1173 (72%) | 265 (55%) | 1655 (70%) | 406 (73%) | 31 (67%) | 114 (77%) | 2353 (72%) | 443 (58%) | 329 (72%) |
| Male | 458 (28%) | 221 (45%) | 720 (30%) | 151 (27%) | 15 (33%) | 35 (23%) | 923 (28%) | 317 (42%) | 126 (28%) |
| **Age [years]** |  |  |  |  |  |  |  |  |  |
| Median (IQR) | 31 (25 - 40) | 33 (26 - 41) | 30 (24 - 38) | 29 (24 - 38) | 31 (26 - 36) | 30 (24 - 36) | 30 (25 - 39) | 33 (27 - 40) | 31 (24 - 36) |
| **WHO clinical stage** |  |  |  |  |  |  |  |  |  |
| I & II | 1046 (64%) | 129 (27%) | 1383 (58%) | 354 (64%) | 17 (37%) | 90 (60%) | 2024 (62%) | 217 (29%) | 250 (55%) |
| III & IV | 585 (36%) | 357 (73%) | 992 (42%) | 203 (36%) | 29 (63%) | 59 (40%) | 1252 (38%) | 543 (71%) | 205 (45%) |
| **Calendar year** |  |  |  |  |  |  |  |  |  |
| Median (IQR) | 2015<br>(2013-2016) | 2012<br>(2010-2013) | 2013<br>(2011-2015) | 2013<br>(2012-2014) | 2013<br>(2011-2013) | 2013<br>(2012-2014) | 2014<br>(2012-2015) | 2012<br>(2010-2014) | 2013<br>(2012-2014) |
| 2005-2013 | 562 (34%) | 368 (76%) | 1256 (53%) | 324 (58%) | 35 (76%) | 83 (56%) | 1438 (44%) | 554 (73%) | 265 (58%) |
| 2014-2016 | 1069 (66%) | 118 (24%) | 1119 (47%) | 233 (42%) | 11 (24%) | 66 (44%) | 1839 (56%) | 207 (27%) | 190 (42%) |
| **CD4 cell count [cells/μL]<sup>&</sup>** |  |  |  |  |  |  |  |  |  |
| Median (IQR) | 301<br>(191 - 434) | 190<br>(90 - 327) | 296<br>(170 - 439) | 308<br>(183 - 447) | 233<br>(124 - 386) | 288<br>(160 - 450) | 299<br>(187 - 438) | 197<br>(100 - 337) | 307<br>(200 - 451) |
| **Follow-up time [months]** |  |  |  |  |  |  |  |  |  |
| Total | 51730 | 9301 | 32663 | 14090 | 1367 | 2692 | 93203 | 16956 | 6904 |
| Median (IQR) | 22 (10 - 43) | 10 (3 - 28) | 6 (1 - 19) | 19 (11 - 33) | 23 (14 - 40) | 9 (3 - 26) | 20 (11 - 39) | 15 (5 - 35) | 8 (2 - 23) |

*\*after correction using tracing data: Outcomes are revised, and all numbers shown are multiplied by inverse probability weights [ML] and averaged over the 50 imputed datasets. For BMA and RIDGE weights, see supplemental Table S1.*
*<sup>&</sup>after imputation of missing values.*

### 3.1 Weights used for correction

For the corrected analyses, the 2117 patients not LTFU were assigned weight 1 and the 1623 patients LTFU and not traced weight 0. Weights of the 752 traced patients depended on the estimation approach used. Supplemental <u>Figure S2</u> shows that distributions of weights were similar amongst the three approaches, and weights highly correlated. Median weights assigned to traced patients were 2.03 (range 1.12-19.68) with ML, 2.47 (range 1.41-15.93) with RIDGE and 2.13 (range 1.16-22.61) with BMA.

### 3.2 Crude incidence of death

<u>Figure 1</u> shows the estimated cumulative probabilities of death, LTFU and being alive after starting ART. Corrected estimates were similar across the three weighting approaches. Compared to the uncorrected estimate of 11.9% (95% CI 10.9-13.0%, Panel A) mortality by four years after starting ART, cumulative mortality doubled in corrected analyses (Panels B to D), ranging from 21.6% (95% CI 18.9-24.6%) to 23.5% (95% CI 20.0-27.6%) depending on the weights used. At the same time, cumulative probability of being alive at 4 years after ART start increased from 29.9% (95% CI 28.3-31.6%) in uncorrected analysis to 63.8% (95% CI 59.6-68.3%) when corrected using ML weights, 65.1% (95% CI 61.7-68.8%) for RIDGE weights and 63.9% (59.8-68.3%) for BMA weights.

**Figure 1:**
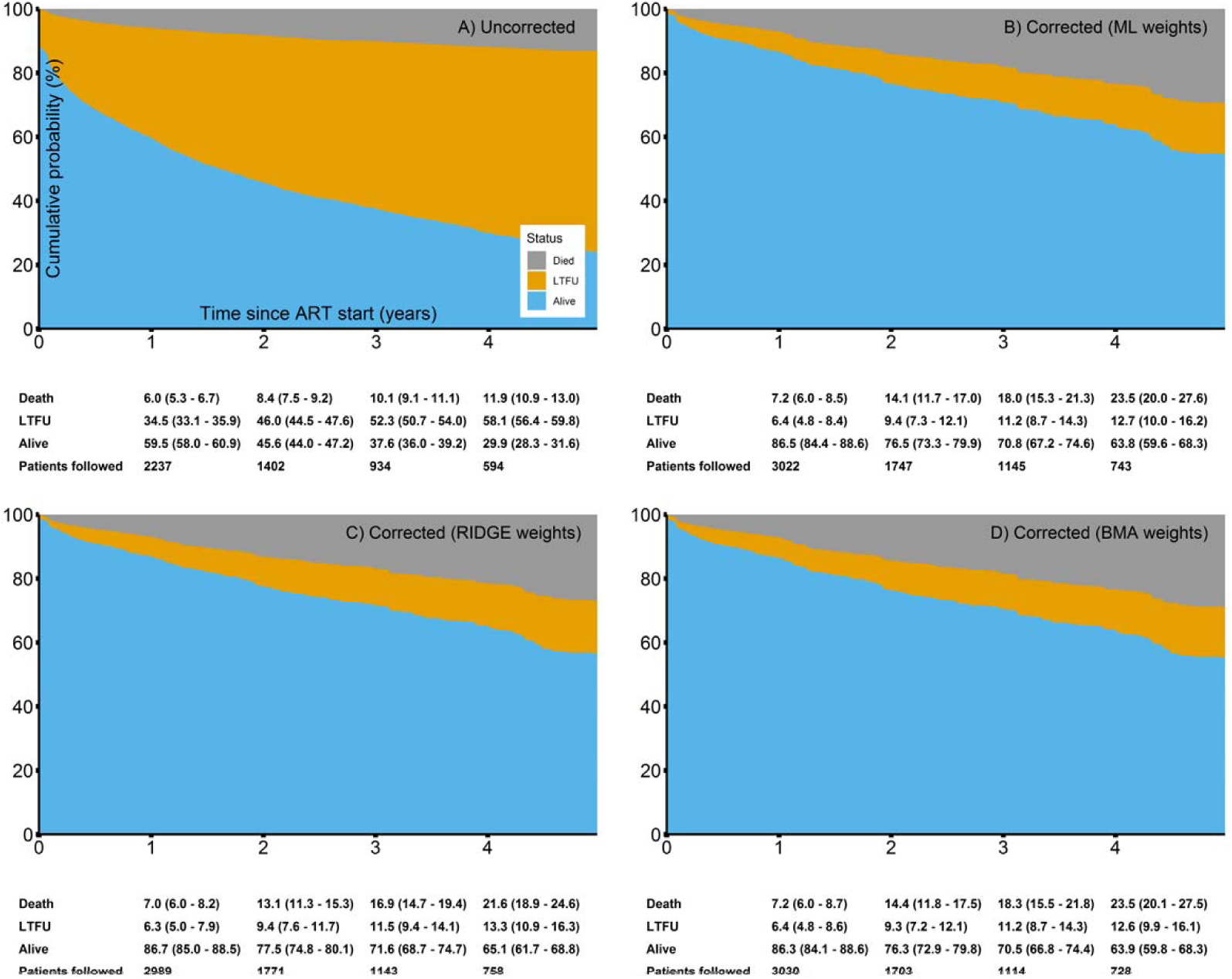
Cumulative incidences of outcomes in patients starting ART, including death, loss to follow-up (LTFU) and being alive. Below the plots, cumulative probabilities (95%-CI) for outcomes at 1-4 years after starting ART are shown. Panel A) reports uncorrected estimats, Panel B)-D) corrected ones (ML weights, RIDGE weights, BMA weights).

### 3.3 Risk factors for death

<u>Figure 2</u> shows uncorrected and corrected adjusted HRs (aHR, adjusted for all other covariates in the model) for mortality. Corrected estimates were similar for all three weighting approaches but differed from uncorrected estimates for some covariates. In the uncorrected analysis, starting ART with lower CD4 counts and more advanced clinical stages was associated with an increased hazard of death. All other associations failed to reach conventional statistical significance. However, there was some evidence for an increased hazard of death with male sex and starting ART in the earlier calendar period.

**Figure 2:**
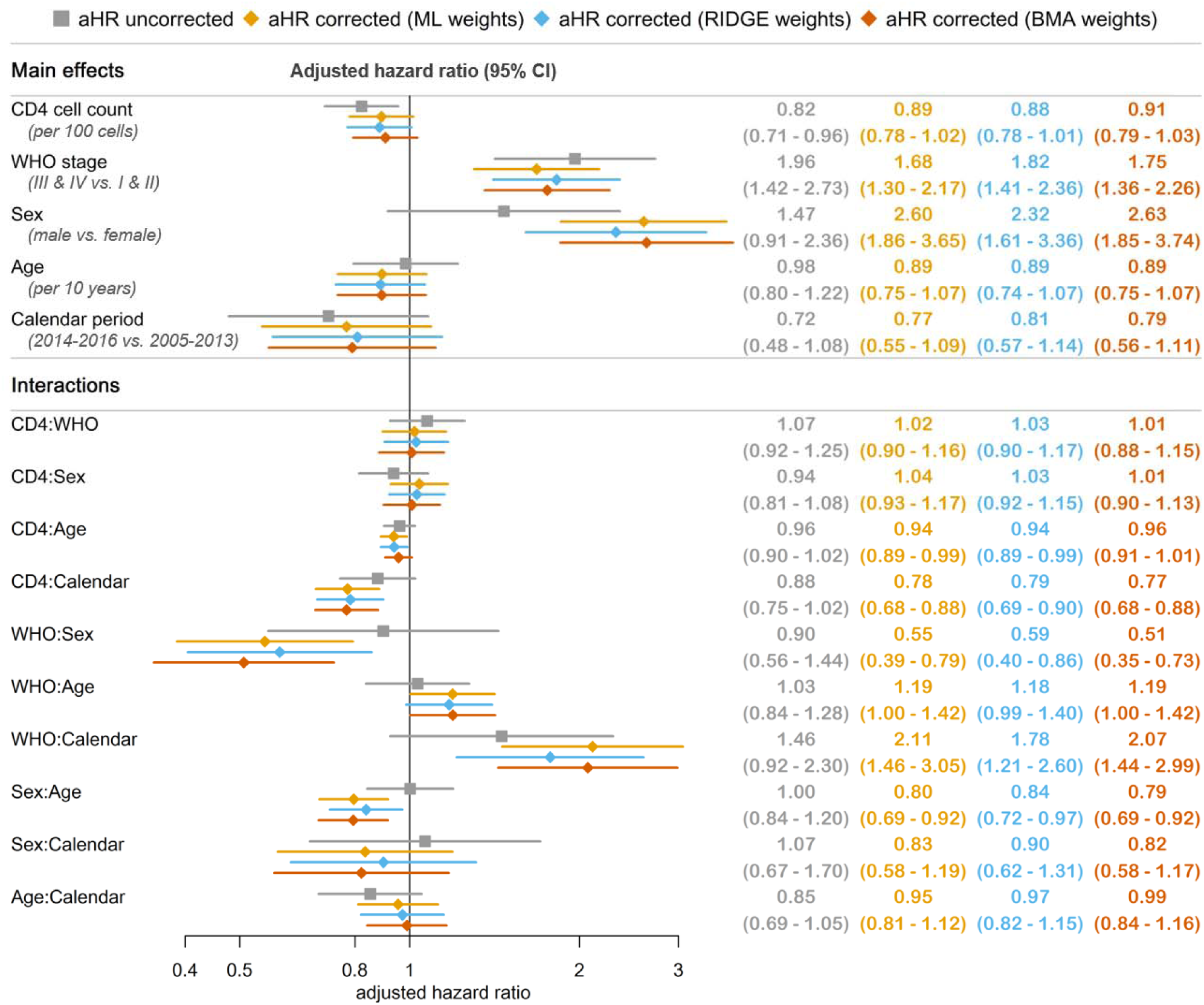
Adjusted hazard ratios (aHR) for outcome death, uncorrected and corrected for true outcomes in patients lost to follow-up.

In corrected analyses, male sex was associated with a substantial increase in the hazard of death, whereas the protective effect of higher CD4 counts became attenuated. Starting ART in more advanced clinical stages was also associated with mortality. The analysis of interactions showed that the latter association was more pronounced in females than males. The association of male sex with mortality became weaker with increasing age. There was also evidence that associations with CD4 count and WHO stage were stronger in later compared to earlier periods. Using ML weights, for example, starting in a more advanced WHO stage was associated with a 68% increase in hazard of death (aHR 1.68) when starting ART 2005-2013, but with a 254% increase when starting ART 2014-2016 (1.68 times interaction coefficient of 2.11).

### 3.4 Risk factors for loss to follow-up

<u>Figure 3</u> shows uncorrected and corrected aHRs for LTFU. In uncorrected analyses, less advanced clinical stages, younger ages and starting ART in later periods was associated with an increased hazard of LTFU. Of note, the association with age was only apparent for patients starting in less advanced clinical stages (stage I & II: aHR 0.87, 95%-CI 0.80-0.95 per 10-year age increase; stage III & IV: aHR 0.97, 95%-CI 0.82-1.16), while the association with calendar period was more pronounced in males (aHR 1.82, 95%-CI 1.30-2.54) compared to females (aHR 1.47, 95%-CI 1.29-1.67). Contrary to uncorrected results, in corrected analyses, there was no evidence for an association with age and the association with WHO stage was only apparent in females (aHR 0.67, 95%-CI 0.49-0.93 comparing stage III & V to stage I & II with ML weights) but not in males (aHR 1.70, 95%-CI 0.71-4.15).

**Figure 3:**
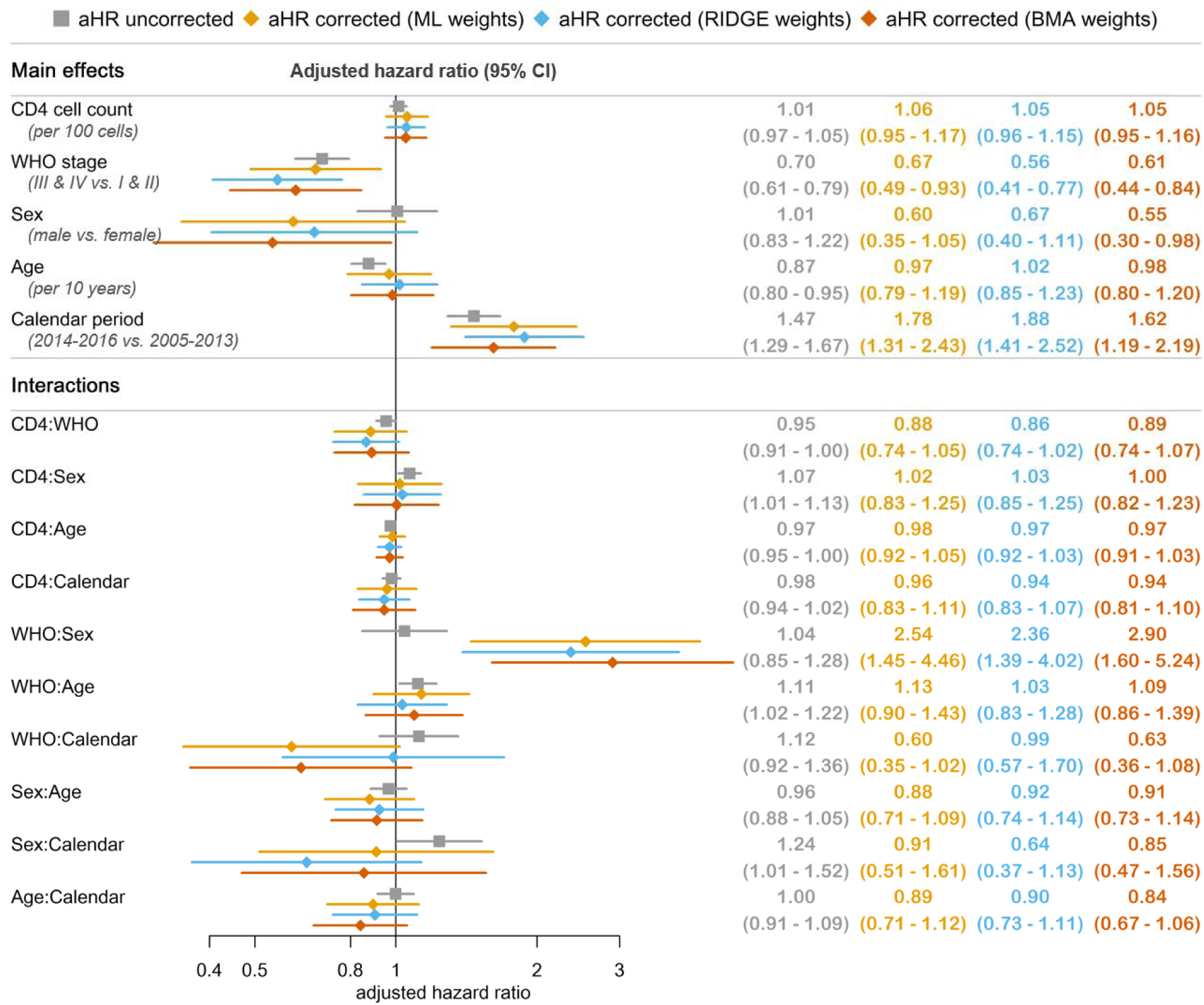
Adjusted hazard ratios (aHR) for loss to follow-up (LTFU), uncorrected and corrected for true outcomes in patients LTFU.

### 3.5 Relative importance of risk factors on cumulative incidence

Supplemental <u>Figure S3</u> shows results from modelling the sub-distribution hazard of death. In the uncorrected analysis, the strongest association (ranked by estimated effect size on the log-scale) was observed for WHO stage, where starting in more advanced stages was associated with an increase in the incidence of death. Second and third strongest associations were seen for calendar period and CD4 count, where starting in earlier periods and with a higher CD4 counts were both associated with a decreased risk of death. All other variables failed to reach conventional statistical significance. In the corrected analysis, male sex was additionally associated with an increased risk of death as well as several interactions between covariates. <u>Figure S4</u> shows corresponding results for the cumulative incidence of LTFU. Results from both corrected and uncorrected analyses corresponded relatively well to the associations obtained with the cause-specific regression model.

### 3.6 Sensitivity Analyses

Supplemental <u>Figures S5-S9</u> show results from analyses restricted to complete cases. Overall, trends were similar to those with imputed CD4 cell counts, but some differences could be obtained.

## 4. Discussion

### 4.1 Main findings

In this ART programme in rural Mozambique, correction using tracing data doubled both estimates of mortality and survival at four years, independently of the type of weights used. Correction also affected some of the associations between patient characteristics and outcomes. For example, corrected analyses showed a higher risk of death in men, which did not reach conventional statistical significance in uncorrected analyses. In addition, both the associations for CD4 count and for WHO stage were modified by calendar period in corrected analysis: lower CD4 counts and more advanced clinical stages increased mortality more pronounced in the later calendar period compared to the earlier one.

### 4.2 Interpretation

The increased hazard of death in men and patients with lower CD4 counts and more advanced clinical stages was expected and in line with results from other studies [9,14,38]. The effect modification by calendar period for CD4 count and WHO stage might be explained by a guideline change: WHO guidelines changed from recommending starting ART at CD4 counts below 350 cells/µL to starting at CD4 counts below 500 cells/µL in 2013. While all patients with CD4 counts between 350 and 500 cells/µL were eligible for ART in the later period, only those who suffered from advanced disease in this group were eligible in the earlier period, thus attenuating these associations.

With regard to the outcome LTFU, results from uncorrected analysis are also very relevant. In uncorrected analyses, “LTFU” represents all patients not returning to the clinic, while in corrected analyses, LTFU represents only patients not returning to the clinic and not found when traced. The increased hazard of LTFU associated with younger ages found in uncorrected analyses is in line with findings from other studies that showed lower adherence in adolescents [39–42]. The increased risk of LTFU associated with less advanced clinical stages reveals that patients who do not return to the clinic might be those who feel well and do not see the need of treatment. These results demonstrate the need to convince young and healthy people of the importance of adhering to treatment.

Other studies have shown the implications of incorporating outcomes of patients LTFU on mortality estimates [9,13,43]. Interestingly, in previous studies, correction using tracing or linkage data increased estimated mortality more substantially than seen in our study. The larger effect may be due to the more modest increase in mortality among the patients who were LTFU and traced in our study, compared to the previous studies, which found higher mortality in patients LTFU [4,6,8]. This discrepancy could be because, in the two clinics included in our analysis, a majority of patients was traced relatively soon after a missed clinic visit. The probability of finding someone alive will, of course, be higher if tracing is done quickly after LTFU [16,43]. We tried to address this by including the duration of being LTFU in the models used to estimate weights. However, the maximum time being lost before traced observed was 4.5 years. As analyses included some patients LTFU with a much longer duration of being lost (and possibly with higher mortality) it might thus be that our results are still underestimating mortality somewhat.

We used inverse-probability weighting (IPW) to correct estimates for true outcomes in patients LTFU. Other studies used different approaches, such as multiple imputation or inflation factors [9,38,43]. Using simulated data, Schomaker and colleagues showed that multiple imputations of survival times performs slightly better than IPW, especially if ascertainment is low. However, the simulated data included only a few covariates and no interactions. As multiple imputation is not well developed for complex survival data and interactions [26], it is questionable if it would outperform IPW methods in real-world settings. Also, our IPW approach probably performs better than methods which do not incorporate tracing or linkage data, or weighting approaches that do not model weights based on patient characteristics.

### 4.3 Strengths and limitations

Few studies of ART outcomes have been done in rural Africa. Because people living in rural areas in Mozambique might differ from those living in urban areas or other countries, results might not be generalisable outside the context of rural Mozambique. Many individuals had missing CD4 counts at ART initiation, which we addressed by multiple imputation. Results based on imputed values were fairly similar to those of complete case analyses, and differences may be explained by the larger proportion of missing CD4 counts amongst patients LTFU. However, if the covariates included in the imputation model do not fully explain differences between patients with and without CD4 cell counts, the missing at random assumption will be violated. Similarly, the patients traced were not selected randomly. The IPW approach is valid if all differences between patients traced and patients not traced are explained by the covariates included in the logistic regression models. If the selection was done randomly, the IPW approach would lead to (approximately) equal weights for all patients traced. This was not the case in our study, indicating that the covariates included in the logistic regression models could explain some of the differences. However, we could still have missed some important covariates that were not collected, such as distance to the clinic. Counsellors may prefer to trace patients that are closer to the clinic, but those who live further away might have a higher risk of death.

The comparison of three different weighting approaches, using maximum likelihood, penalised maximum likelihood and Bayesian model averaging is a strength of this study. Weights were highly correlated and, unsurprisingly, results similar with very few exceptions. This demonstrates, on the one hand, the usefulness of applying different correction methods to the same data to assess the stability of results. On the other hand, it also indicates that all three estimation methods might be suitable to correct mortality estimates for true outcomes in patients LTFU (if all important covariates explaining differences between patients traced and not traced are collected).

### 4.4 Conclusions

In conclusion, our study showed the importance of data on outcomes in patients LTFU to correct clinic-level estimates. The corrected estimate of cumulative mortality differed substantially from the uncorrected one, and correction had some impact on associations between patient characteristics and the hazard of death. Tracing studies are urgently needed not only from a clinical point of view to bring patients back into care but also for epidemiological and programmatic reasons to estimate outcomes of ART programmes accurately.

## Authorship contribution statement

**Nanina Anderegg:** Conceptualisation, Methodology, Formal analysis, Visualisation, Writing – original draft, Writing – review & editing. **Jonas Hector:** Conceptualisation, Data curation, Resources, Writing – review & editing. **Laura F Jefferys:** Data curation, Resources, Writing – review & editing. **Juan Burgos:** Data curation, Resources, Writing – review & editing. **Michael A Hobbins:** Resources, Supervision, Writing – review & editing. **Jochen Ehmer:** Resources, Supervision, Writing – review & editing. **Lukas Meier:** Methodology, Supervision, Writing – review & editing. **Marloes H Maathuis:** Methodology, Supervision, Writing – review & editing. **Matthias Egger:** Conceptualisation, Supervision, Supervision, Funding acquisition, Writing – original draft, Writing – review & editing.

### Declarations of interes

None

## Data Availability

The datasets generated during the current study and analysed are not publicly available but are available from the corresponding author on reasonable request.

## Acknowledgements

The research reported in this publication was supported by the National Institute of Allergy and Infectious Diseases of the National Institutes of Health under Award Number U01AI069924. The content is solely the responsibility of the authors and does not necessarily represent the official views of the National Institutes of Health. ME was supported by special project funding (grant 17481) from the Swiss National Science Foundation.

